# The genetics and epidemiology of *N-* and *O-* IgA glycomics

**DOI:** 10.1101/2024.01.05.24300885

**Authors:** Alessia Visconti, Niccolò Rossi, Albert Bondt, Agnes Hipgrave Ederveen, Gaurav Thareja, Carolien A. M. Koeleman, Nisha Stephan, Anna Halama, Hannah J. Lomax-Browne, Matthew C. Pickering, Xu-jie Zhou, Manfred Wuhrer, Karsten Suhre, Mario Falchi

**Author notes:** These authors contributed equally (Alessia Visconti, Niccolò Rossi). These authors share senior authorship (Manfred Wuhrer, Karsten Suhre, Mario Falchi).

## Abstract

Immunoglobulin (Ig) glycosylation modulates the immune response, and plays a critical role in ageing and diseases. Studies have mainly focused on IgG glycosylation, and little is known about the genetics and epidemiology of IgA glycosylation. Here, we generated, using a novel LC-MS method, the first large-scale IgA glycomics dataset in serum from 2,423 twins, encompassing 71 *N-* and *O-*glycan species. We showed that, despite the lack of a direct genetic template, glycosylation is highly heritable, and that glycopeptide structures are sex-specific, and undergo substantial changes with ageing. We observe extensive correlations between the IgA and IgG glycomes, and, exploiting the twin design, show that they are predominantly influenced by shared genetic factors. A genome-wide association study identified eight loci associated with both the IgA and IgG glycomes (*ST6GAL1*, *ELL2*, *B4GALT1*, *ABCF2*, *TMEM121*, *SLC38A10*, *SMARCB1*, *MGAT3*), and two novel loci specifically modulating IgA *O-*glycosylation (*C1GALT1* and *ST3GAL1*). Validation of our findings in an independent cohort of 320 individuals from Qatar showed that the underlying genetic architecture is conserved across ethnicities. Our study delineates the genetic landscape of IgA glycosylation and provides novel potential functional links with the aetiology of complex immune diseases, including genetic factors involved in IgA nephropathy risk.

## Introduction

Immunoglobulins are large molecules produced by plasma cells and activated B cells in response to exposure to antigens. Immunoglobulins (Ig) undergo complex co- and post-translational modifications including the selective addition of oligosaccharides (glycans) to the side chains of asparagine (*N*-glycosylation), serine or threonine (*O*-glycosylation) residues in the peptide backbone. These modifications, despite being non-template driven, are genetically determined^1,2^. So far, IgG has attracted the interest of most of the epidemiological and genetic research in Ig glycobiology, with studies showing that IgG glycosylation affects effector functions^3^ with relevant implications in ageing and disease, including, among others, autoimmune and infectious diseases, and cancer^3^. Conversely, less is known about the genetics and epidemiology of the IgA glycome. IgA is the most abundantly secreted immunoglobulin in the body, the second most prevalent antibody isotype in blood after IgG, and is heavily glycosylated^1–4^. Notably, while the CH2 domain of each of the four IgG subclasses (IgG_1-4_) contains a single *N-*glycosylation site^5^, the two IgA subclasses (IgA_1-2_) carry multiple conserved *N*-glycosylation sites^5^. In addition, IgA_1_ can also undergo *O*-glycosylation^4^. Aberrant *O*-glycosylation has been observed in cancer and autoimmunity^6^, suggesting a role for both IgA *O*- and *N*-glycosylation in health and disease. Abnormally glycosylated IgA1 has been found to be the driving pathogenic force in IgA nephropathy, the most common form of glomerulonephritis worldwide. Here, we explore the genetic architecture of the circulating IgA *O*- and *N*-glycome, and compare glycosylation features between the IgA and IgG molecules to assess the impact of shared genetics and environment on these differently specialised antibody isotypes.

## Results

### Cohort description

Study subjects were 2,423 individuals of European ancestry from the TwinsUK cohort^7^ (513 monozygotic and 615 dizygotic twin pairs, and 167 singletons), mainly females (78%), between 19 and 88 (mean=58, standard deviation (SD) = 11) years old (**Supplementary Table 1**). Using LC-MS, we quantified 33 *O*- linked and 38 *N*-linked IgA_1-2_ glycan species at the glycopeptide level^8^. Their names are here composed of the letter codes of the first three amino acids of the tryptic peptide sequence (**Figure 1a, Supplementary Table 2**). Additionally, we summarised glycosylation features (*e.g.*, bisection, fucosylation, galactosylation, sialylation) across structurally related *O*- and *N*- glycans expressed on the same glycopeptide into 7 and 21 derived traits, respectively (**Supplementary Table 3**). In the same sample, we also quantified 36 IgG_1-4_ *N*- linked glycans and calculated 16 IgG_1-4_ derived traits^9^ (**Supplementary Tables 2** and **3**). Derived traits provide a composite and more robust measure of Ig glycosylation than single measured glycopeptides^9^ and will be the main focus of this work.

**Figure 1.**
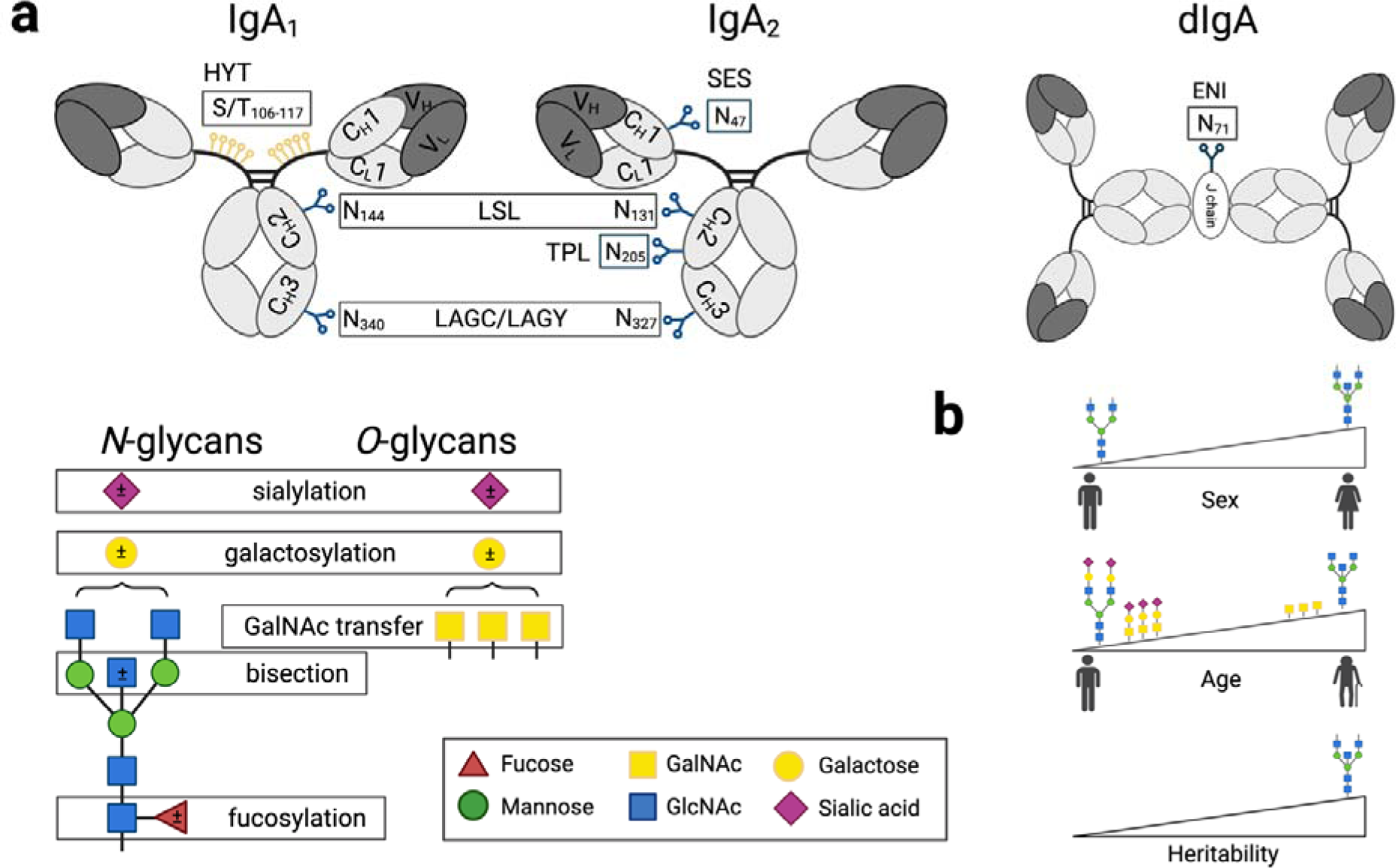
(**a**) **Schematic representation of IgA_1_ and IgA_2_ with examples for *O-* and *N*-glycan structures.** *N*-glycans are attached to the amino side chain of an asparagine residue (N), while *O*-glycans are attached to the side chain of serine (S) or threonine (T). Each IgA_1_ heavy chain contains two *N-*glycosylation sites (*i.e.*, at N144 and N340), while the hinge region has six *O*-glycosylation sites (*i.e.*, at T106, T109, S111, S113, T114, and T117), all present on a single tryptic peptide. IgA_2_ has four *N-*glycosylation sites on the heavy chain (*i.e.*, N47, N131, N205, and N327), plus one additional *N-*glycosylation site at the J-chain (N71). The three-letter code defines the tryptic (glyco-)peptides: HYT for the *O*-glycosylation at the hinge region of IgA_1_; LSL for the *N*-glycosylated sites N144 or N131 on IgA_1_ or IgA_2_, respectively; LAG for the *N*-glycosylated sites N340 or N327 on IgA_1_ or IgA_2_, respectively, which were detected with either a terminal tyrosine (LAGY) or as the truncated form (LAGC); SES for the *N*-glycosylated site N47 on IgA_2_; TPL for the *N*-glycosylated site N205 on IgA_2_; ENI for the *N*-glycosylated site N71 at the J-chain on IgA_2_. Glycosylation site numbering is according to Uni-ProtKB. Monosaccharides symbols and example structures of *O*- and *N*-glycans and of derived traits are also shown. (**b**) **Visual summary of the heritability of the IgA glycome and of its association with sex and age**. Bisected *N*-glycans have higher abundance in females. Ageing is associated with increased *N*-glycan bisection, and decreased *N*- and *O*-glycan sialylation and galactosylation. Bisected *N*-glycans are also the most heritable traits.

### IgA glycosylation is sex-specific and undergoes substantial changes with ageing

We found that IgA *N*-glycan bisection, regardless of the glycopeptide on which it was expressed, was higher in females (*P*≤4.53×10^-9^; **Figure 1b, Supplementary Table 4 and 5**). Age was significantly associated with 68% of the IgA derived traits (**Supplementary Tables 6 and 7**), indicating increased *N*-glycan bisection and decreased *N*- and *O*-glycan sialylation and galactosylation with ageing (**Figure 1b**). IgG glycosylation data showed concordant results, consistent with previous reports^10^ (**Supplementary Tables 6 and 7**).

### IgA glycosylation is highly hereditable

Using the classical twin model, we showed IgA glycosylation to be significantly heritable, with derived traits displaying mean h^2^=50.8% (SD=13.8%; max h^2^=74.3%; **Figure 2, Supplementary Table 8 and 9**), similarly to IgG derived traits (mean h^2^=52.2%, SD=12.1%, max h^2^=71.0%; **Supplementary Figure 1, Supplementary Table 8 and 9**). Bisected *N*-glycans were among the highest heritable traits in both isotypes. A shared environmental component was estimated to contribute (mean c^2^=21.3%; SD=2.2%) to the variance of sialylation and galactosylation of diantennary IgA *N-*glycans at the LSL and SES glycopeptides (**Figure 2, Supplementary Table 8 and 9**). Interestingly, IgA measured *O*-glycans where overall more influenced by environmental factors than *N*-glycans (Wilcoxon P=3.29 ×10^-4^).

**Figure 2.**
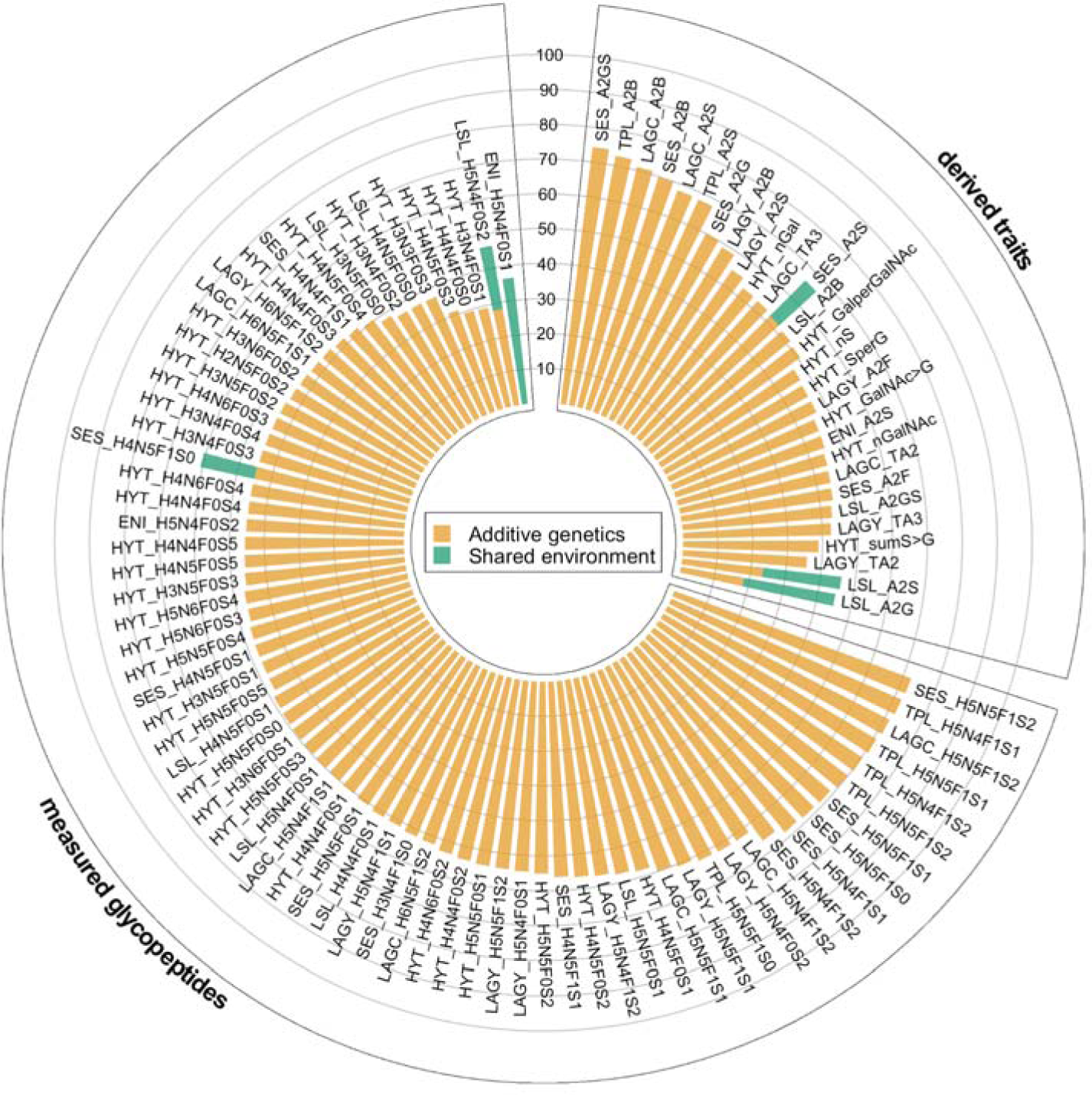
Heritability of IgA measured glycopeptides and derived traits. We used the ACE model to partition the variance for each trait into additive genetic (orange), and shared (green) and unique (white) environmental components.

### IgA and IgG glycomes show extensive genetic correlation

IgA measured glycopeptides were significantly correlated (**Supplementary Table 10**), particularly those expressed on the same glycopeptide, as expected, since most of them represented substrates and products of enzymatic reactions. Interestingly, using their ratios as proxies for enzymatic reaction rates, we observed a consistent reduction in enzymatic activity along with growing substrate complexity for progressive *N*- acetylgalactosaminylation, sialylation, and galactosylation of IgA_1_ *O*-glycan structures (*P*<2.2×10^-16^; **Figure 3a**; **Supplementary Figures 2-4**), possibly due to either decreased substrate availability or increased steric hindrance. Consistent results were observed for IgA and IgG *N*-glycans (**Supplementary Figures 3 and 5**). IgA derived traits were also significantly correlated among each other (**Figure 3c, Supplementary Table 11**), but, more interesting, 69 pairwise correlations were observed among IgA and IgG derived traits at a Bonferroni-corrected threshold of P<2.2×10^-4^ and at |P|>0.25, suggesting that shared genetic and/or environmental factors can simultaneously influence the behaviour of different antibody isotypes (**Supplementary Table 12**). Using a bivariate variance component model, we showed these correlations between IgA and IgG derived traits to be mostly explained by shared genetic factors (median shared heritability=79%, IQR=73%-83%; **Supplementary Table 13**), with shared environmental factors also playing a significant role (median contribution 21%; IQR=17%-27%). The largest shared genetic component was observed for those pairs of IgA-IgG derived traits describing *N*-glycans sialylation and bisection (**Figure 3b**), the latter also being the most heritable derived trait in both IgA and IgG.

**Figure 3.**
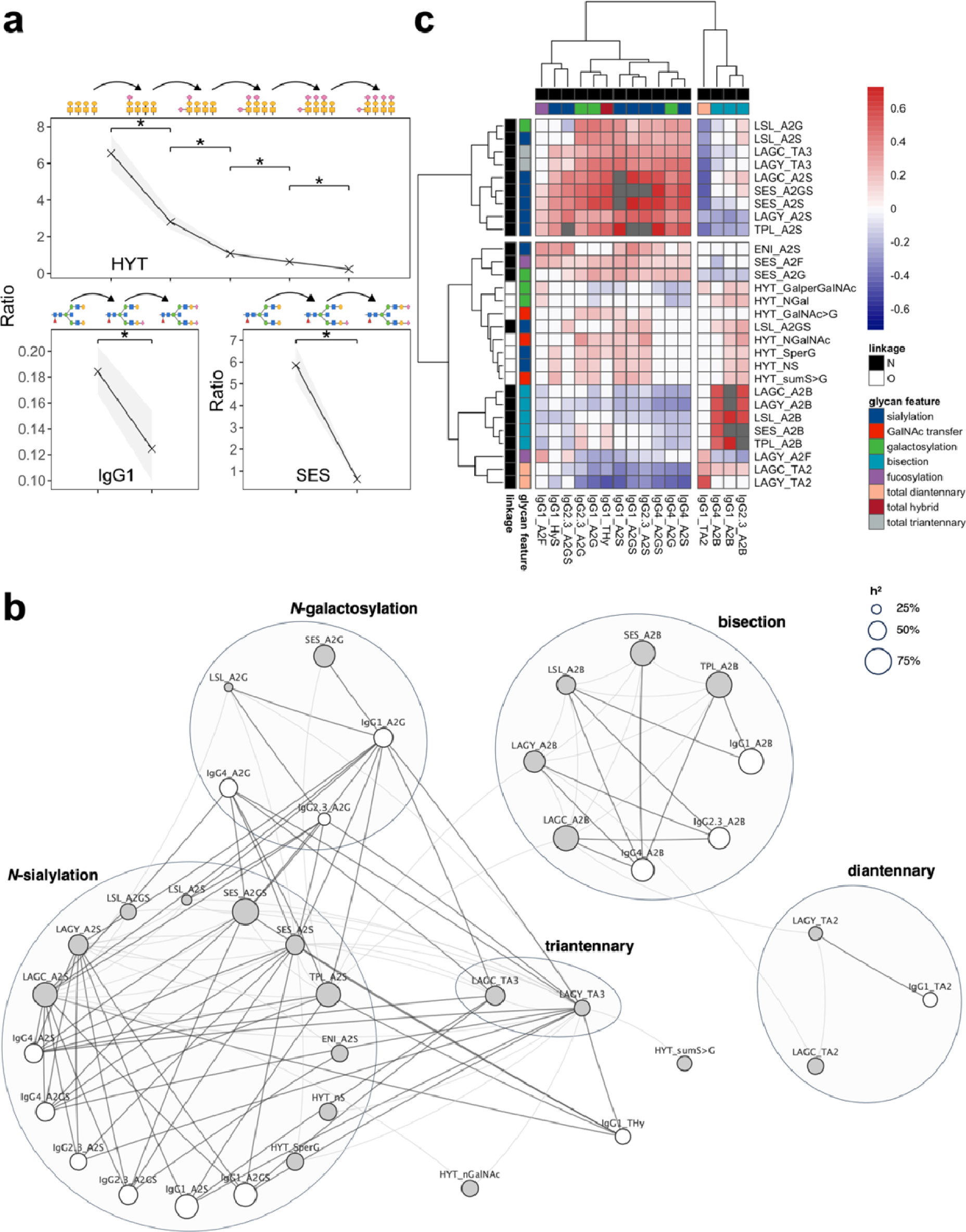
**(a) Progressive decrease in sialylation efficiency of different *N*- and *O*-glycan structures**. Crosses represent the median value of the ratio of the glycan structures depicted on top of the panel at the corresponding position of the x-axis. These glycan structures differ by a single sialic acid residue, and thus reflect sequential sialylation reactions in the glycosylation pathway. The grey area shows the interquartile range of each ratio. Significant differences, evaluated by means of the Wilcoxon test, are indicated with an asterisk (*P*<2.2×10^-16^). (**b**) **Correlation network of IgA and IgG *N*-linked glycosylation derived traits.** Each node represents a derived trait (IgG: white; IgA: grey), with size proportional to the trait heritability. Edges connect phenotypically correlated (Pearson’s |P|>0.25) derived traits. IgA-IgG correlations are shown with darker, thicker lines compared IgA-IgA correlations. For IgA, only correlations between derived traits on different peptides are shown. Derived traits are grouped according to their glycosylation features. (**c**) **Genetic correlation of IgA and IgG derived traits.** The heatmap shows IgA derived traits in rows and IgG derived traits in columns, with colour labels indicating the linkage (*O-* or *N-*) and the glycosylation feature (*e.g.*, sialylation, fucosylation, bisection). Cells colours represent the genetic correlation coefficients with grey cells indicating IgA-IgG pairs whose calculation did not converge. Derived traits are hierarchically clustered based on absolute genetic correlation coefficients using the *hclust* function as implemented in the pheatmap R package (v1.0.12).

### Genetics of the IgA glycome

To identify genes involved in this shared heritability and explore the genetic architecture of the serum IgA glycome, we performed a genome-wide association study (GWAS) of IgA glycosylation in 2,371 individuals at 5,411,460 common (minor allele frequency, MAF>5%) single nucleotide polymorphism (SNPs), using a linear mixed model to account for non-independence of observations within twin pairs, and adjusting for sex, age, LC-MS plate number, and the first five principal components of the genotype data. The average genomic inflation factor (λ_GC_) was 1.01 (max=1.04), thus suggesting that there was no residual confounding by population stratification nor cryptic relatedness, nor any apparent systematic genotyping error. At a Bonferroni-corrected threshold of *P*<1.35×10^-9^ (**Methods**), we identified 87 genome-wide significant associations of 45 measured glycopeptides and 17 derived traits at 11 genetic loci (**Figure 4, Table 1, Supplementary Table 14, Supplementary Data 1**). Conditional association analyses identified no further independent genome-wide significant associations. Lead SNPs explained, on average, 5% of the phenotypic variance of the associated IgA traits (IQR=3%-6%, calculated on measured glycopeptides and derived traits) and 9% of their genetic variance (IQR=6%-13%; **Supplementary Tables 15 and 16**). Novel genetic associations for *O*-glycan traits were identified at the genes encoding for the glycosylation enzymes core 1 synthase, glycoprotein-N-acetylgalactosamine 3-β-galactosyltransferase (C1GALT1) and ST3 β-Galactoside L-2,3-Sialyltransferase 1 (ST3GAL1), and were replicated in an independent multi-ethnic cohort of 320 individuals from Qatar (QMDiab^11,12^; **Supplementary Tables 1**) at a conservative Bonferroni-corrected threshold of *P*<6.94×10^−4^ (**Methods; Table 1 and Supplementary Table 14**). The lead SNP rs10246303-T at the *C1GALT1* locus tagged reported associations for decreased modified Tiffeneau-Pinelli index^20^, with an association for chronic obstructive pulmonary disease (COPD) being reported within the same locus^21^ (**Table 1, Supplementary Table 14**). A third newly identified locus for *O*-glycans mapping to the gene encoding for the glycosylation enzyme polypeptide N-Acetylgalactosaminyltransferase 12 (GALNT12) showed comparable effects between TwinsUK and QMDiab, but did not reach statistical significance in the replication, likely due to the low frequency (MAF<1.5%) of the associated SNPs in QMDiab (MAF_TwinsUK_>7%; **Supplementary Table 14**).

**Figure 4.**
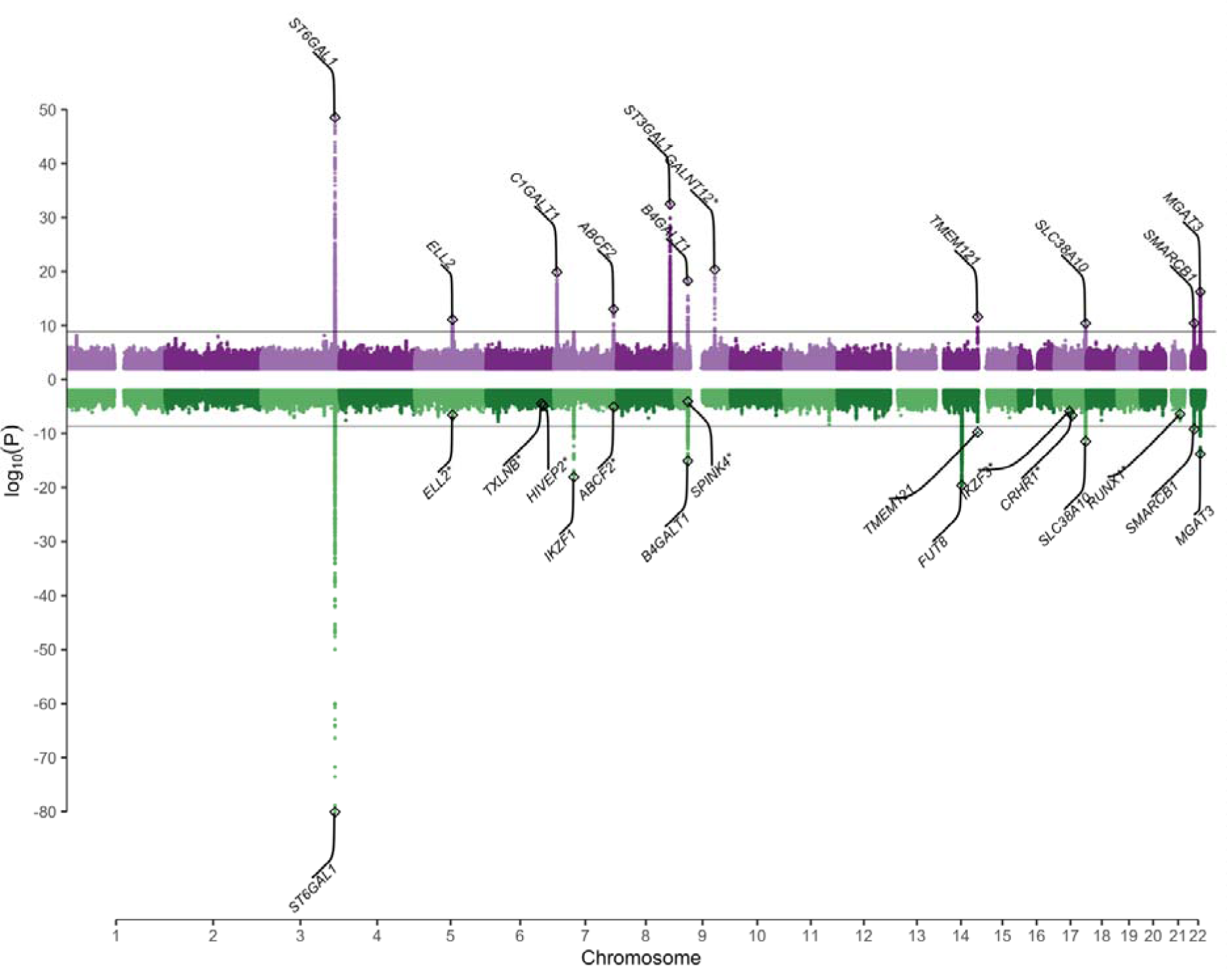
Miami plot showing the genome-wide association of IgA (top) and IgG measured glycopeptides and derived traits (bottom). The x-axis shows the genomic coordinates (GRCh37.p13) of the tested SNPs and the y-axis shows the –log10 P value of their association. The horizontal black line indicates the threshold for genome-wide significance at 1.35×10^−9^ and 2.08×10^−9^, for IgA and IgG, respectively. Asterisks indicate newly identified IgA glycosylation loci failing replication in QMDiab (top panel) and replicated IgG loci^3^ not reaching GWAS-significance in the present study (bottom panel).

**Table 1.**
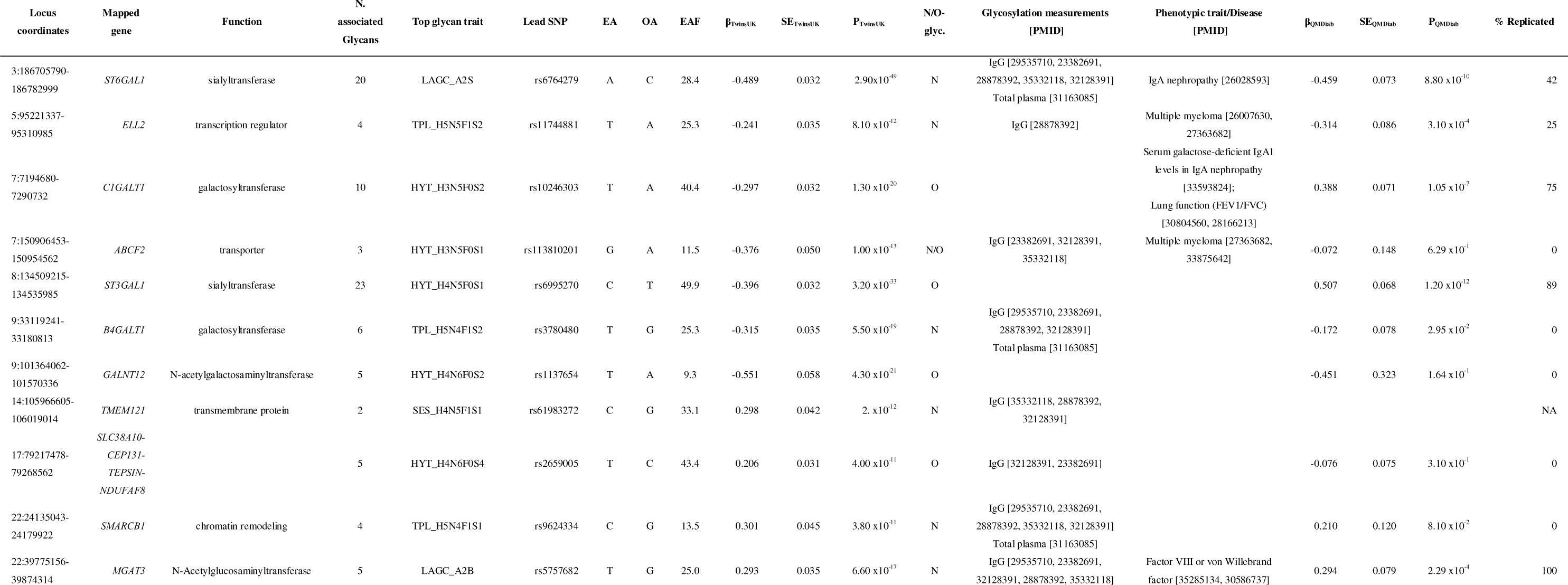
Results of the IgA GWAS. For each locus, the table reports the cytogenic location, the candidate gene along with its function, the total number of associated glycan traits (either measured glycopeptides or derived trait), as well as the fraction, expressed as a percentage, of replicated glycan associations in QMDiab (% Replicated), the strongest associated SNP, along with the effect allele (EA, the minor allele), the other allele (OA), and the effect allele frequency (EAF) in TwinsUK. For each lead SNP within the locus, we report the glycan trait showing the strongest association, along with its GWAS summary statistics, *i.e.*, the association effect size (β), standard error (SE) and p-value (P) in both TwinsUK and QMDiab. We further report whether the locus is associated with either *N*- or *O*-glycans in TwinsUK (N/O glyc), as well as information on previous GWAS (identified as [PMID]) reporting associations within the locus for the *N*-glycome (Glycosylation measurements) or relevant traits or disease (Phenotypic trait/Disease), as extracted from the GWAS Catalog.

At the remaining eight loci identified, associations for *O*-glycans (*ABCF2* and *SLC38A10*) and *N*-glycans (*ABCF2*, *B4GALT1*, *ELL2*, *MGAT3*, *ST6GAL1*, *SMARCB1*, and *TMEM121*) were in strong linkage disequilibrium (LD) (r^2^>0.8) with GWAS associations previously reported for *N*-glycan traits^2^ (**Table 1 and Supplementary Table 14**). IgA *N*-glycans associations at the *ST6GAL1*, *MGAT3*, and *ELL2* loci were also identified in QMDiab (**Table 1 and Supplementary Table 14**), with lead SNPs at the *ST6GAL1* and *ELL2* loci being coincident or in strong linkage disequilibrium (LD) with previously reported associations for IgA nephropathy and multiple myeloma risk, respectively (**Table 1**).

### Shared variants influence both IgA and IgG glycosylation

GWAS of the IgG *N-*glycan traits replicated previous associations (*P*<7.72×10^−5^; **Methods**; **Figure 4**; **Supplementary Table 18**) at 16 out of the 27 loci identified by the largest (n=8,090) GWAS published to date using a ultra-high-performance liquid chromatography (UHPLC)-measured IgG *N*-glycome^2^, with eight loci reaching genome-wide significance (*P*<2.08×10^-9^; **Supplementary Tables 18 and 19, Supplementary Data 2**), thus suggesting good cross-platform reproducibility. Lead SNPs explained, on average, 5% of phenotypic variance of the associated IgG measured glycopeptides and derived traits (IQR=2%-6%, calculated on measured glycopeptides and derived traits) and 8% of their genetic variance (IQR=4%-10%; **Supplementary Tables 15 and 16**).

Our GWAS highlighted coincident associations between IgA and IgG glycosylation at eight loci, and 23 out of the 69 pairs of matching IgA/IgG derived *N*-glycan traits under significant shared genetic control showed coincident association at the *ST6GAL1*, *B4GALT1*, *MGAT3*, and *SMARCB1* genes. The Bayesian bivariate approach implemented in GWAS-PW^13^ suggested that the same genetic variants were influencing both IgA and IgG correlated traits (posterior probability of model 3>0.90; **Supplementary Table 20**). Overall, all the genetic variants identified for the 69 genetically correlated IgA-IgG pairs explained only a minor fraction of the estimated genetic correlation (median=6%, IQR=0.4%-10%; **Supplementary Table 21**), thus suggesting that a much larger sample size may be needed and/or that the shared genetic effects cannot be fully captured by common SNPs.

## Discussions

IgA is the most abundantly secreted immunoglobulin in the body, and it is heavily glycosylated^4^. Yet, little is known about its genetic architecture, and the shared genetic and environment factors which shape both the IgA and IgG molecules. Here, we used here a high-throughput LC-MS method to generate the first large-scale dataset for the simultaneous analysis of IgA and IgG glycopeptides in a large cohort of twins from the UK.

We showed that bisected *N-*glycans have higher abundance in females. possibly reflecting a sex-specific modulation of immune response, with increased bisecting *N*-acetylglucosamine abundances of IgG in females (identified also in this study) previously observed from childhood^14^. Additionally, we showed that ageing is associated with increased *N-*glycan bisection, and decreased *N-* and *O-*glycan sialylation and galactosylation, consistent with previous findings on IgG^10^. Increased abundances of bisection and, mostly, reduced sialylation and galactosylation on IgG *N-*linked glycopeptides have been connected to several inflammatory and autoimmune conditions, and to ageing^3^.

Exploiting the twin design, we identified extensive genetic correlations between the IgA and IgG glycomes, with bisected *N-*glycans being the most heritable traits in both IgA and IgG. The largest shared genetic component was observed for those pairs of IgA-IgG derived traits describing *N*-glycans sialylation and bisection, the latter also being the most heritable derived trait in both IgA and IgG. Our genome-wide association of the IgA glycome identified ten associated genetic loci, including two novel loci involved in *O*-glycosylation (*C1GALT1* and *ST3GAL1*) and eight loci affecting the abundance of both IgA and IgG glycans (*ST6GAL1*, *ELL2*, *B4GALT1*, *ABCF2*, *TMEM121*, *SLC38A10*, *SMARCB1*, *MGAT3*). These results were validated using 320 individuals of Arab, South Asian, and Filipino descent, showing that the underlying genetic architecture is conserved across ancestries.

Coincident associations at the loci encompassing the *C1GALT1* and *ST6GAL1* genes contribute to a better understanding of the factors involved in IgA nephropathy (IgAN) risk. Galactose deficiency of IgA_1_ *O*- glycans in the hinge region (gd-IgA) has been repeatedly observed in IgAN^15^, with serum gd-IgA being traditionally assessed using ELISA. Association for higher serum gd-IgA levels have been reported at rs10238682-G^16^, a positive regulator of *C1GALT1* expression, genome-wide significant in this study and in strong LD (*R*^2^>0.8) with our lead SNP at the *C1GALT1* gene. C1GALT1 catalyses the transfer of galactose to *N-*acetylgalactosamine *O*- linked to the hydroxy group of threonine or serine residues. We find here that SNPs at this locus associate with increased galactosylation (HYT_nGal and HYT_GalperGalNAc), and lower proportion of degalactosylated acetylgalactosamine relative to galactose (HYT_nGalNAc>nG) in the hinge region. In a previous high-resolution glycomics case-control study of IgA in IgAN susceptibility carried out by our team, we found that these three derived traits were strongly correlated with ELISA measurements of gd-IgA, but they were not associated with IgAN risk or kidney function^8^. Indeed, we suggested that *N-* and *O-*sialylation, rather than *O*-galactosylation, were likely involved in the pathophysiology of IgAN. In this context, the *ST6GAL1* gene, which encodes a sialyltransferase that adds sialic acid to the terminal galactose of *N-*glycoproteins, has been associated with IgAN risk in 8,313 cases and 19,680 controls from China^17^, as well as with disease severity and progression^18^. The associated SNP rs7634389 (r^2^>0.8 with our lead SNPs at *ST6GAL1*), associates with decreased *ST6GAL1* expression, and, here, with lower *N-*sialylation at most derived traits (LAGC_A2S, LAGY_A2S, LSL_A2GS, SES_A2GS, SES_A2S, TPL_A2S), overlapping glycan traits associating with IgAN in our previous study and confirming the involvement of *N-*sialylation in disease risk. Additionally, we validated, at a suggestive Bonferroni-derived threshold of 0.05/25=0.002, three loci reported by a recent large (10,146 cases and 28,751 controls) multi-ancestry GWAS of IgAN^19^, including the risk allele rs10896045-A at the *OVOL1/RELA* gene associating in our study to decreased galactosylation and consequent decreased sialylation at O-glycans (*P*=8.7×10^-4^ at HYT_nS>nG; **Supplementary Table 17**).

Associations for decreased modified Tiffeneau-Pinelli index^20^ and COPD^21^ were reported at the *C1GALT1* locus. IgA is the most prevalent Ig in the lungs, and is altered in chronic respiratory disease accompanied by a decreased pulmonary capacity, including COPD^22^. However, little is known about the similarity between IgA glycosylation in the lung mucosa and in serum, with considerable differences already been observed between circulating and salivary IgA glycosylation^23^.

Associations for increased serum IgA levels^24^ and lower multiple myeloma risk^25,26^ have been reported at rs3815768-T and rs56219066-C, respectively, in LD (*R*^2^>0.8) with our lead SNP rs11744881-T at *ELL2*, here associated with lower IgA *N*-glycan sialylation. Increased sialylation is found in IgG paraproteins from patients with multiple myeloma^27^, where it correlates with metastatic behaviour^28^. As ELL2 influences both secretory-specific Ig production and *N*-glycan biosynthesis^29^, it is suggested to increase the risk of malignant transformation by reducing Ig levels, which may result in slower antigen clearance and prolonged B-cell stimulation^30^, possibly in a context of aberrantly glycosylated immunoglobulins. Intriguingly, while *ELL2* associates also with IgG sialylation^1^, the multiple myeloma risk allele rs56219066-C shows stronger association with circulating IgA levels compared to IgG^26^, suggesting that IgA, more than IgG, *N*-glycan sialylation may play a role in multiple myeloma.

In conclusion, we show here that serum IgA *N*- and *O*-glycosylation is largely genetically determined, and that the underlying genetic architecture is conserved across ancestries. Ageing is accompanied by increased bisection and decreased *N*- and *O*-glycan sialylation and galactosylation, with bisection being also the most heritable trait. We report profound parallels between IgA and IgG *N*-glycosylation, suggesting shared genetic and environmental mechanisms of immune regulation to simultaneously orchestrate different specialised antibody isotypes. We identified common genetic mechanisms of immune regulation between IgA and IgG *N*-glycans. However, the major fraction of the shared IgA-IgG glycome heritability remains unexplained, and the shared environmental determinants remain to be explored. Taken together, these findings expand our understanding of the architecture of circulating Ig glycosylation and provide potential functional link with the aetiology of complex diseases, including of factors involved in IgAN risk.

## Methods

### Discovery Cohort

Study subjects were 2,423 individuals of self-declared European ancestry from the TwinsUK cohort^7^ (**Supplementary Table 1**). TwinsUK includes about 14,000 monozygotic and dizygotic twin volunteers from all regions across the United Kingdom, unselected for any disease and representative of the general population. This study was carried out under TwinsUK BioBank ethics, approved by Northwest – Liverpool Central Research Ethics Committee (REC reference 19/NW/0187), IRAS ID 258513.

### Replication Cohort

The Qatar Metabolomics Study on Diabetes (QMDiab) is a cross-sectional case–control study of diabetes with 374 participants of Arab, South Asian, and Filipino descent^11,12^ (**Supplementary Table 1**). For 320 samples joint genetics and glycomics data were available. All study participants were enrolled between February 2012 and June 2012 at the Dermatology Department of HMC in Doha, Qatar. The initial study was approved by the institutional review boards of Hamad Medical Corporation (HMC) and Weill Cornell Medicine - Qatar (WCM-Q) (research protocol #11131/11). Written informed consent was obtained from all participants.

### Mass Spectrometric Glycomics

IgA and IgG glycans were quantified in serum samples using LC-MS, as previously described^8,23^. In total, 33 IgA_1_ *O*-linked glycopeptides, 38 IgA_1-2_ *N*-linked glycopeptides, and 36 IgG_1-4_ *N*-linked glycopeptides were retained and quantified. The absolute signal intensities were normalised to the intensity sum of all glycopeptide species sharing the same tryptic peptide sequence, resulting in relative abundances. In this manuscript, IgA_1_ and IgA_2_ glycopeptide names are composed of the letter codes of the first three amino acids of the peptide sequence: HYT, LSL, TPL, SES, ENI, and LAG, the last detected in two variants (*i.e.*, as LAGC and LAGY). The peptide name is followed by the glycan composition indicating the number of hexoses (H), *N-*acetylhexosamines (N), fucoses (F), and sialic acids (S; **Figure 1a, Supplemental Table 2**). Structurally similar glycopeptides were summarised into derived traits calculated from their relative intensities (**Supplemental Table 3**), resulting in 7 IgA_1_ *O*-glycan, 21 IgA_1-2_ *N*-glycan, and 16 IgG_1-4_ *N*- glycan derived traits. Because each measured glycopeptide structure carries different types of monosaccharides, derived traits can give a more composite and robust measure of the different glycosylation features. Outliers, defined as measurements deviating more than four standard deviations from the mean of each trait, were removed. To ensure the normality of their distribution, measured glycopeptides and derived traits were quantile normalised.

### Association with age and sex

Association of measured glycopeptides and derived traits with age at sample collection and sex was assessed by fitting a linear mixed model in the R statistical framework (*lmer* function, lme4 package v1.1-31) with plate number and family structure modelled as random effects. We used the approach proposed by Li & Ji^31^ to determine the effective number of independent tests to control for the family-wise error rate, resulting in 37 and 24 tests, for IgA and IgG, respectively. Associations were considered significant if the p-value was lower thanL0.05/37 = 1.35×10^-3^ and 0.05/24 = 2.08×10^-3^ for IgA and IgG, respectively.

### Heritability estimation in TwinsUK

We used the *mets* R package^32^ (v1.2.5) to estimate the contribution of additive genetic (A), shared (C) and individual-specific environment (E) effects on measured glycopeptides and derived traits variations (ACE model), using a classical twin design study. The ACE model was then compared with the most parsimonious AE model, which did not include the effect of the common environment, and the CE and E model, which hypothesise that the trait variability only under environmental control. Models were compared through Akaike’s information criterion (AIC). Quantile-normalised measured glycopeptides and derived traits were corrected for plate number (included as random effect in a linear mixed model, *lmer* R function), and passed to *mets*. Sex and age at the sample collection were included as covariates. Four pairs of monozygotic twins and three pairs of dizygotic twins were removed because the twins were either adopted or reared apart.

### IgA correlations

Pairwise IgA measured glycopeptides and derived traits correlation was estimated using Pearson product– moment correlation on pre-processed glycan abundances. More in detail, quantile-normalised measured glycopeptides and derived traits were adjusted for age, sex (fixed effects), plate number, and family structure (random effects) using a linear mixed model (*lmer* R function), and the correlations were computed on the residuals. Associations were considered significant if the absolute correlation coefficient was greater than 0.25 and the p-value was lower thanL0.05/(37×36/2) = 7.51×10^-5^, where (37×36/2) corresponds to the number of elements of a lower triangular matrix with size 37×37, and 37 corresponds to the effective number of independent tests as described above.

### IgA glycan ratios

Ratios were calculated using untransformed relative frequencies of pairs of measured glycopeptides expressed on the same peptide, after outliers were removed. For *N*-glycans, we selected IgA and IgG measured glycopeptides representing the product–substrate of known sialylation and galactosylation reactions in the Ig *N*-glycosylation pathway^33^. For *O*-glycans, reactions of *N*-acetylgalactosaminylation, sialylation, and galactosylation were estimated based on the ratios of pairs of measured *O-*glycopeptides which differed for a single GalNAc, sialic acid and galactose residue, respectively. Overall, 21 multi-step enzymatic conversions were identified, involving at least two enzymatic transformations of the same underlying glycan structure, as exemplified below:

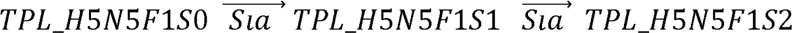

Differences in the distribution of ratios representing two consecutive glycosylation reactions were assessed using the Wilcoxon test, and p-values less than 0.05/31 = 2.38×10^-3^, were considered significant, where 31 corresponds to the number of comparisons performed.

### IgA-IgG correlations and shared heritability

Pairwise additive genetic (ρG) and environmental (ρE) correlations among IgA-IgG derived traits were estimated through bivariate maximum likelihood-based variance decomposition in SOLAR-Eclipse^34^ (v8.1.1; http://www.solar-eclipse-genetics.org/). Age and sex were included as covariates. SOLAR-Eclipse was also used to estimate the phenotypic correlations (ρP) so that the same proportion of variance associated with covariates is removed for phenotypic and genetic correlation estimates. Likelihood ratio tests were used to assess whether ρP, ρG, and ρE were significantly different from zero. Phenotypic correlations were considered significant if the absolute correlation coefficient was larger than 0.25 and the associated p-value was lower than 0.05/(19×12)=2.19×10^-4^, where 19 and 12 correspond, respectively, to the effective number of independent IgA and IgG derived traits tested estimated as described above.

For pairs of correlated IgA-IgG glycan traits, we estimated the genetic (COV_G_) and environmental (COV_E_) covariances based on the following formulas:

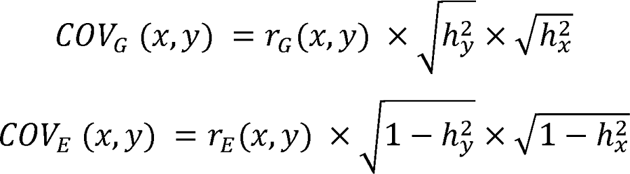

where *h^2^_x_* and *h^2^* are the heritabilities (h^2^) of traits x and y, as estimated by SOLAR-Eclipse, and *r_G_* (*x*, *y*) and *r_E_* (*x*, *y*) are the genetic and environmental correlations, respectively. Genetic/environmental correlation coefficients were constrained to zero if the associated p-value was larger than 0.05, so that the resulting covariance is zero. Then, the proportion of traits phenotypic correlation explained by genetics (*i.e.*, shared heritability) was estimated based on the following equation:

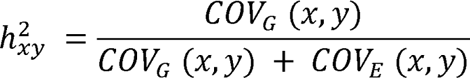

which assumes that the total phenotypic covariance is given by the sum of the genetic and environmental covariances COV_G_ and COV_E_, respectively.

### Genotyping

In TwinsUK, microarray genotyping was conducted using a combination of Illumina arrays (HumanHap300, HumanHap610, 1M-Duo and 1.2M-Duo 1LM) and imputation was performed using the Michigan Imputation Server^35^ using haplotype information from the Haplotype Reference Consortium (HRC) panel^36^ (r1.1). Genotypes were available for 2,371 individuals with glycomics data. A total of 5,411,460 SNPs meeting the following conditions were included in our genome-wide association study: call rate ≥95%, MAF>5%, and imputation score >0.4.

In QMDiab, genotyping was carried out using Illumina Omni array 2.5 (v8). Standard quality control of genotyped data was applied, with SNPs filtered by sample call rateL>98%; SNP call rate >98%, and Hardy-Weinberg equilibrium (HWE) P<1×10^−6^. Imputation was done using SHAPEIT software with 1000G phase 3 version 5 and mapped to the GRCh37 human genome build. Imputed SNPs were filtered by imputation quality >0.7.

### GWAS discovery step

To take into account the non-independence of the twin data, the association with IgA and IgG measured glycopeptides and derived traits in the discovery cohort was performed using GEMMA^37^ (v0.98.1), assuming an additive genetic model and including sex, age at the sample collection, the first five principal components assessed on the genomic data, and plate number as covariates. Associations were considered significant and taken forward for replication if their discovery p-value was lower than 5×10^−8^/37=1.35×10^−9^ and 5×10^−8^/24=2.08×10^−9^, for IgA and IgG, respectively.

### Identification of independent signals within loci

We used a stepwise procedure to identify independent signals within the loci identified in the discovery cohort. For each locus, we fitted a new regression model, where the top-associated genome-wide significant SNP was included as a covariate (*i.e.*, conditional model). Genome-wide significant (*P*<5×10^−8^) SNP resulting from the conditional model was considered as an independent signal and was included in the covariate set of a new conditional model. We stopped the stepwise procedure when we could not identify any additional genome-wide significant SNPs. Conditional models were built using GEMMA^37^ (v0.98.1).

### Replication step

We attempted replication of independent signals for IgA measured glycopeptides and derived traits using data from the QMDiab cohort. Replication was evaluated at the lead SNP of each locus, or at a tag SNP in high linkage disequilibrium (R^2^≥0.8, distance ≤500kb) with any genome-wide significant SNP within the locus. The LD structure was assessed with LDlink^38^ (v3.6) using the available GBR population from 1000 genomes project phase 3. Genetic association was conducted using a linear regression model adjusting for age, sex, diabetes status, and the first three genetic principal components, and was considered replicated if the direction of the effects was concordant between the two cohorts, and if the p-value was lower thanL0.05/72=6.94×10^−3^, where 72 is the number of IgA measured glycopeptides and derived traits-genomic locus pair identified in the discovery step.

To assess the replicability of loci reported by previous GWAS for serum IgG *N*-glycans, we retrieved the reported lead SNPs for the 27 independent genome-wide significant loci identified by the largest (n=8,090) GWAS published to date on circulating IgG glycome^2^. We considered an association replicated in TwinsUK if the p-value of the association for any IgG measured glycopeptides and derived traits at the lead SNP of each locus was lower than 0.05/(27×24)=7.72×10^-5^, where 27 is the number of loci tested and 24 is the effective number of IgG measured glycopeptides and derived traits tested.

### Identification of known loci

We interrogated the NHGRI-EBI GWAS Catalog^39^ (release 2023-02-03, association *P*<5×10^−8^) to identify previously reported associations in coincidence or in strong linkage disequilibrium with those identified and replicated in our study. Additionally, we queried our results to identify associations in coincidence or in strong linkage disequilibrium with 25 genome-wide significant loci reported by a large IgAN meta-analysis^19^, and passing a Bonferroni-derived threshold of 0.05/25=0.002. Linkage disequilibrium was assessed using LDlink^38^ with the parameters listed above.

### Variants annotation

Locus-gene mapping was performed using OpenTarget (https://genetics.opentargets.org/, release 22.10, October 2022). OpenTarget used the “locus-to-gene” (L2G) model to prioritise likely causal genes. An L2G score is derived for a given SNP–gene pair by aggregating evidence from different mapping strategies, including physical distance from the transcription start site of nearby genes, splicing (sQTL) and expression (eQTL) quantitative trait loci colocalization, and chromatin interaction mapping, as well as *in silico* predictions of SNP functional consequence.

### Shared genetic effect

For each pair of derived traits for which a replicated genome-wide significant signal was identified, we applied a Bayesian bivariate analysis, as implemented in the GWAS-PW software^13^ to investigate the presence of a shared genetic effect in the TwinsUK cohort. Briefly, GWAS-PW estimates the posterior probability of quasi-independent genomic regions to include a genetic variant which *(i)* associated only with the first derived trait (model 1), *(ii)* associated only with the second derived trait (model 2), *(iii)* associated with both derived traits (model 3) or *(iv)* that the genomic block includes two genetic variants, associating independently with each of the two derived traits (model 4). We defined regions for which a genome-wide significant signal was identified for at least one of the two traits and showed a posterior probability larger than 0.9 for model 3 or model 4 as characterised by an underlying shared genetic effect or by colocalization of effects, respectively.

Quasi-independent regions (n=1,703) were pre-determined by estimating LD blocks in European populations^16^. Since our GWASs were performed on overlapping sets of individuals, we estimated the expected correlation in the effect sizes from the GWAS summary statistics using *fgwas*^40^, as suggested in^13^, and used this correction factor in the GWAS-PW modelling.

### SNP-based heritability

For a given SNP*_i_*, we calculated the proportion of explained phenotypic variance in TwinsUK as:

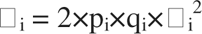

where β*_i_* is the estimated effect of SNP*_i_* in univariate analysis, and *p_i_* and *q_i_*are the minor and major allele frequencies of SNP*_i_* respectively, as estimated in TwinsUK. For measured glycopeptides and derived traits that had more than one independent genome-wide significant association, we estimated the total SNP-based variance explained as the sum of the proportion of variance explained by each of the top independent SNPs.

### GWAS signals contribution to IgA-IgG shared heritability

To evaluate the contribution of GWAS signals to IgG-IgA glycome shared heritability, we estimated pairwise IgA-IgG derived traits additive genetic covariance upon conditioning on the lead SNPs in TwinsUK. More in detail, for each pair tested, derived traits were independently adjusted for age, sex, plate number, and lead SNPs at genome-wide-significant loci associated with any of the two traits (*lmer* R function). Standardised residuals were then passed to SOLAR-Eclipse^34^ (v8.1.1) to estimate the conditional phenotypic and genetic correlations. The additive genetic covariance was estimated based on the following formula:

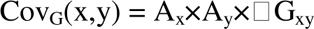

where *A_x_*and *A_y_* are the square roots of the heritabilities (h^2^) of traits x and y, and ρ*G_xy_* is the additive genetic correlation.

## Data availability

Data generated during the study are available as Supplementary Data. Data on TwinsUK twin participants are available to bona fide researchers under managed access due to governance and ethical constraints. Raw data should be requested via our website (http://twinsuk.ac.uk/resources-for-researchers/access-our-data/) and requests are reviewed by the TwinsUK Resource Executive Committee (TREC) regularly. Consent of QMDiab participants does not include public posting of genomics data.

## Acknowledgements

TwinsUK is funded by the Wellcome Trust, Medical Research Council, European Union, Chronic Disease Research Foundation (CDRF), and the National Institute for Health Research (NIHR)-funded BioResource, Clinical Research Facility and Biomedical Research Centre based at Guy’s and St Thomas’ NHS Foundation Trust in partnership with King’s College London. The authors acknowledge the use of the research computing facility at King’s College London, Rosalind (https://rosalind.kcl.ac.uk) and King’s Computational Research, Engineering and Technology Environment (CREATE, https://docs.er.kcl.ac.uk/).

KS was supported by the Biomedical Research Program at Weill Cornell Medicine in Qatar, a program funded by the Qatar Foundation. We thank the staff of the HMC dermatology department and of WCM-Q for their contribution to QMDiab. Finally, we are grateful to all study participants for their invaluable contributions to this study.

## Authors contributions

MF, MW, and KS conceptualised and designed the project. MW devised and supervised the mass spectrometric glycomics. AB performed the mass spectrometric glycomics with assistance from AHE. CAMK performed the mass spectrometric glycomics on the QMDiab cohort. AV and NR carried out the statistical analyses with contributions from MF. NS and AH were responsible for the QMDiab data collection and management. GT performed the imputation of the QMDiab genotypes. KS performed the GWAS replication in the QMDiab cohort. AV, NR, KS, MW, and MF interpreted the results with contributions from HJLB and MCP. AV, NR, and MF wrote the manuscript with contributions from XZ, KS, and MW. All authors approved the final version of the manuscript.

