## Supplementary Figures for "The genetics and epidemiology of *N-* and *O-* IgA glycomics"

*Shared genetic factors shape N- and O-glycosylation patterns of circulating immunoglobulins A and G*

[†]These authors contributed equally

[#]These authors share senior authorship

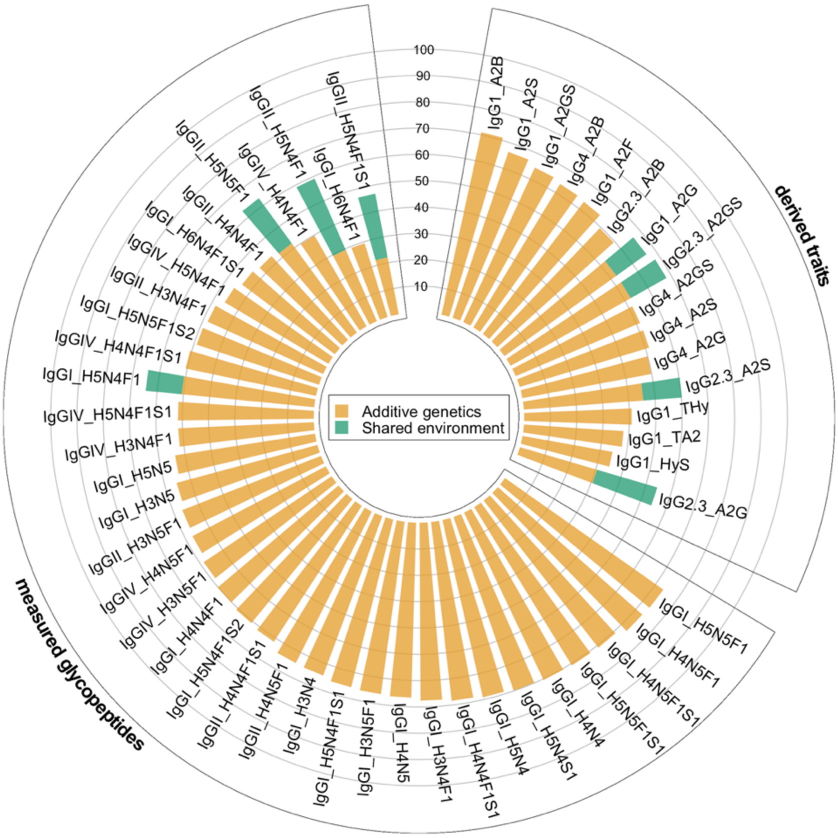


**Supplementary Figure 1**. **Heritability of IgG measured glycopeptides and derived traits.** We used the ACE model to partition the variance for each glycan trait into additive genetic (orange), and shared (green) and unique (white) environmental components.

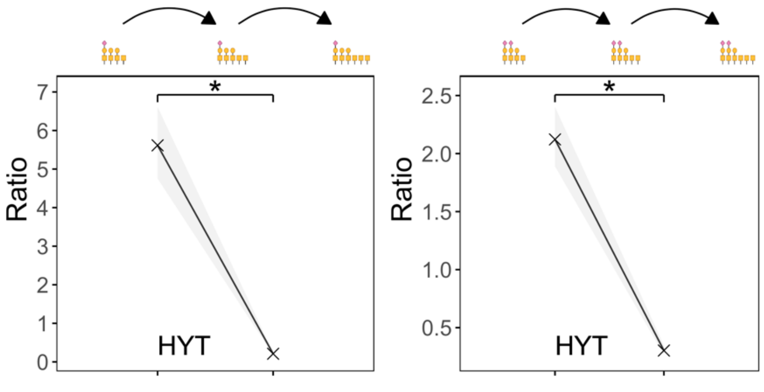


**Supplementary Figure 2. Progressive decrease in GalNAc transfer efficiency of two different growing *O*-glycan structures on IgA1.** Crosses represent the median value of the ratio of the glycan structures depicted on top of the panel at the corresponding position of the x-axis. These glycan structures differ for a single GalNAc residue, and thus reflect sequential transfer of GalNAc residues in the *O*-glycosylation pathway. The grey area shows the interquartile range of each ratio. Significative differences, evaluated by means of the Wilcoxon test, are indicated with an asterisk ($P<2.2\times10^{-16}$)

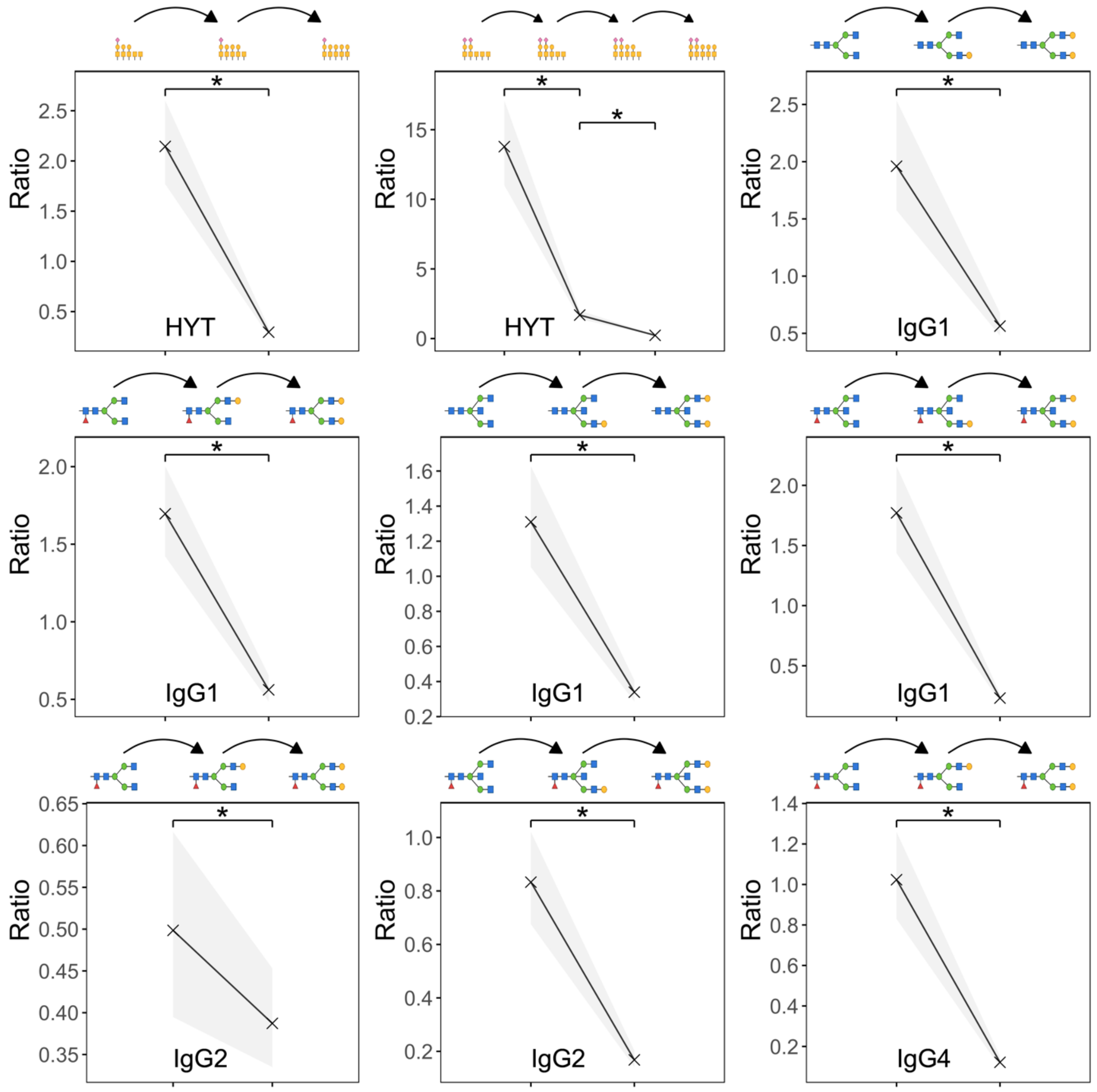


**Supplementary Figure 3. Progressive decrease in galactosylation efficiency of growing *O*- and *N*-glycan structures on IgA and IgG, respectively.** Crosses represent the median value of the ratio of the glycan structures depicted on top of the panel at the corresponding position of the x-axis. These glycan structures differ for a single galactose residue, and thus reflect sequential galactosylation reactions in the glycosylation pathway. The grey area shows the interquartile range of each ratio. Significative differences, evaluated by means of the Wilcoxon test, are indicated with an asterisk ($P<2.2\times10^{-16}$).

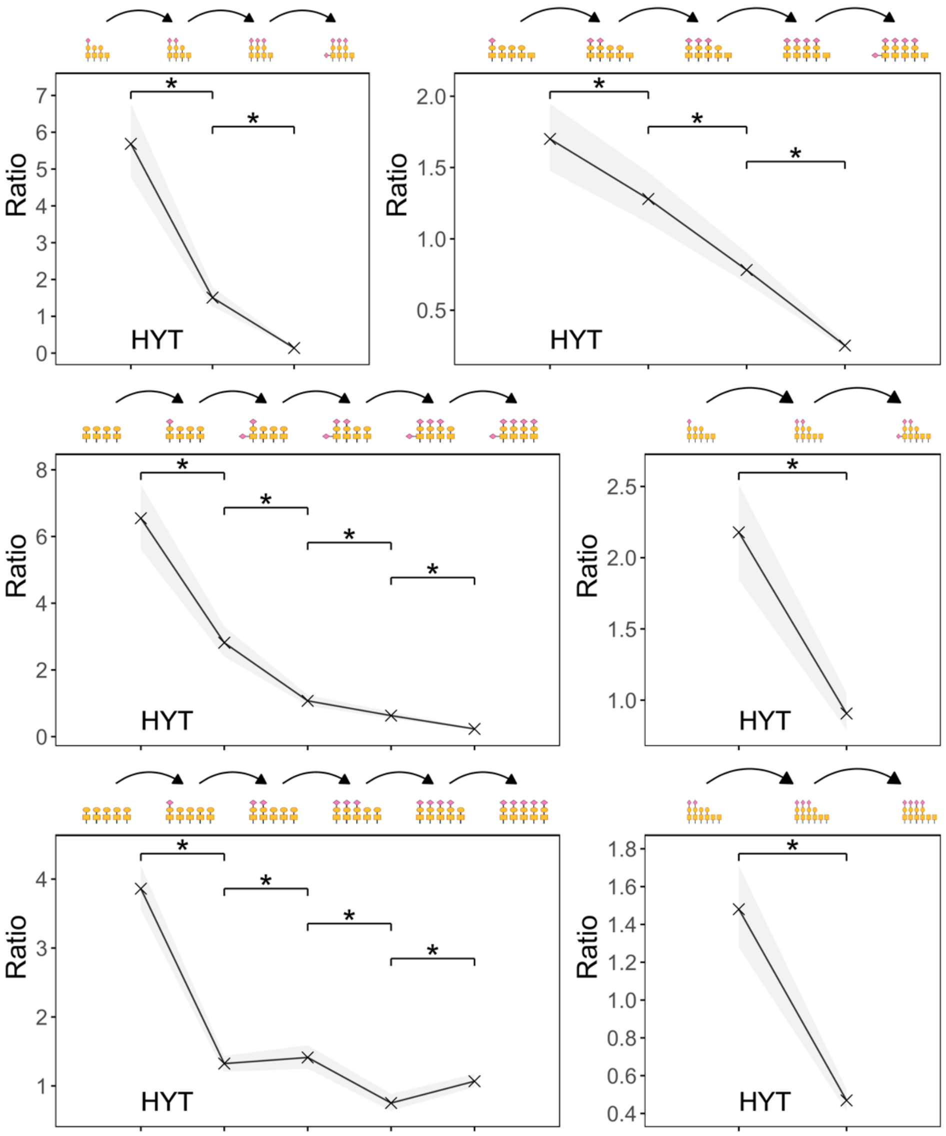


**Supplementary Figure 4. Progressive decrease in sialylation efficiency of growing *O*-glycan structures on $IgA_1$.** Crosses represent the median value of the ratio of the glycan structures depicted on top of the panel at the corresponding position of the x-axis. These glycan structures differ for a single sialic acid residue, and thus reflect sequential sialylation reactions in the *O*-glycosylation pathway. The grey area shows the interquartile range of each ratio. Significative differences, evaluated by means of the Wilcoxon test, are indicated with an asterisk ($P<2.2\times10^{-16}$).

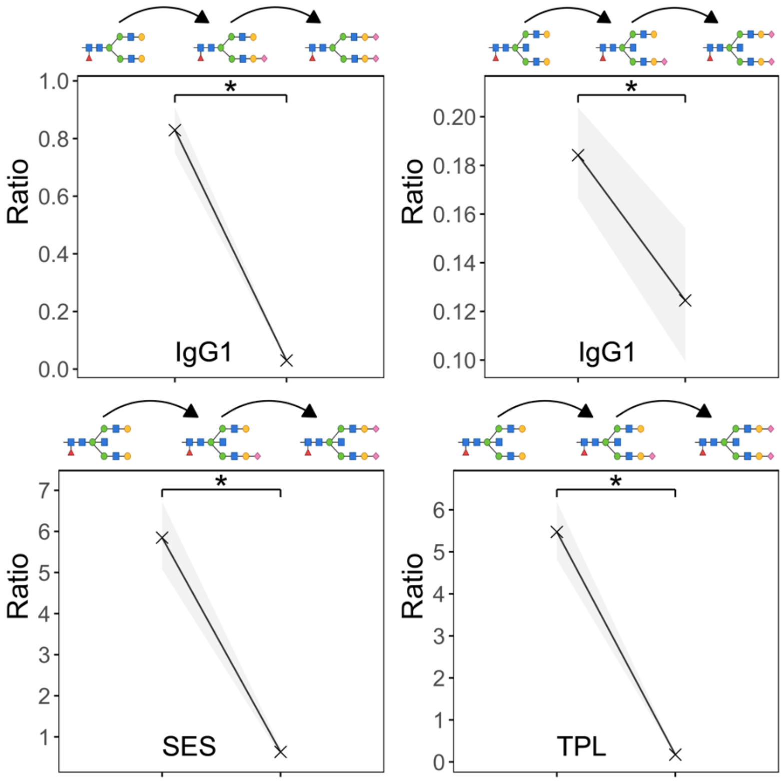


**Supplementary Figure 5. Progressive decrease in sialylation efficiency of growing *N*-glycan structures on IgA and IgG.** Crosses represent the median value of the ratio of the glycan structures depicted on top of the panel at the corresponding position of the x-axis. These glycan structures differ for a single sialic acid residue, and thus reflect sequential sialylation reactions in the *N*-glycosylation pathway. The grey area shows the interquartile range of each ratio. Significative differences, evaluated by means of the Wilcoxon test, are indicated with an asterisk ($P<2.2\times10^{-16}$).
